# Informative Presence Bias in Comorbidity Data of Medicare Advantage-Enrolled Beneficiaries

**DOI:** 10.1101/2024.01.16.24301389

**Authors:** Justin M. Schaffer, Austin Kluis, John J. Squiers, Jasjit K. Banwait, Mario F. L. Gaudino, Michael J. Mack, J. Michael DiMaio

**Author notes:** ADDRESS FOR CORRESPONDENCE Justin Schaffer Baylor Scott and White - The Heart Hospital, Plano 4708 Alliance Blvd. Plano, TX 75093.

## Abstract

**Background:** Analyses of Medicare administrative claims data are faced with methodological challenges, including accounting for the potential effect of insurance status on documented comorbidities. We present an example of how failing to account for informative presence bias related to beneficiary enrollment status in such analyses may lead to flawed results.

**Methods:** In this retrospective observational study of Medicare beneficiaries undergoing isolated coronary artery bypass grafting (CABG) from 1999-2019, we compare the distribution of documented comorbidities between beneficiaries with Medicare Advantage (MA) and Traditional Medicare (TM) plans. Long-term survival was then compared in both unweighted and overlap weighted analyses with and without the inclusion of documented comorbidities.

**Results:** Among 3,015,066 Medicare beneficiaries undergoing CABG from 1999-2019, 2,345,476 underwent isolated CABG and had suitable data for analysis. The annual proportion of MA-enrolled beneficiaries undergoing CABG remained stable from 1999-2007 (1.1-4.5%) and then progressively increased annually, reaching 38.2% in 2019. The incidences of documented comorbidities were substantially lower among MA-enrolled versus TM-enrolled beneficiaries. Among MA-enrolled and TM-enrolled beneficiaries, respectively, the unweighted median survival difference was only 8 [-12,28] days (10.02 [9.96,10.07] vs 10.00 [9.98,10.01] years); the weighted (adjusted for demographics and procedural characteristics, but not beneficiary comorbidities) median survival difference was also minimal at -2 [-28,24] days (10.00 [9.95,10.06] vs 10.01 [9.98,10.04] years). However, the weighted (with adjustments including beneficiary comorbidities) median survival difference demonstrated a substantial survival disadvantage for MA-enrolled beneficiaries compared to their TM-enrolled counterparts: -604 [-626,-575] days (9.78 [9.73,9.83] vs 11.44 [11.41,11.47] years), respectively.

**Conclusions and Relevance:** Comorbidities in MA-enrolled beneficiaries may be severely under-reported in Medicare data. Studies failing to account for this are susceptible to informative presence bias with a significant treatment effect. In the absence of policy changes, increasing enrollment in MA plans will continue to decrease the population of Medicare beneficiaries with suitable data for study in comparative analyses.

## INTRODUCTION

The Centers for Medicare and Medicaid Services (CMS) maintains a database of administrative claims that is increasingly used in clinical research.^1^ Hospital claims submitted to CMS using International Classification of Diseases clinical modification (ICD-CM) and procedural (ICD-PCS) codes are available in the Medicare Provider Analysis and Review (MedPAR) file for analysis by research groups.^2^ CMS also maintains an annual record of beneficiary demographics in the Master Beneficiary Summary File (MBSF). The chronic conditions segment of the MBSF (CC-MBSF) identifies individual beneficiary status (and date of initial diagnosis) for chronic conditions and has become a standard tool to adjudicate comorbidities in Medicare analyses.^3^

Studies using Medicare claims data are contingent upon the correct interpretation of coding data to characterize beneficiary comorbidities, procedural characteristics, and outcomes.^4^ In 2015, CMS transitioned from ICD-9 to ICD-10 coding; studies spanning this transition must accurately handle the coding change.^5^ Importantly, CC-MBSF data has been adjudicated across the ICD-9/10 transition, making it suitable for use in such analyses.^6^ It has been noted, however, that beneficiaries who enroll in Medicare Advantage (MA) plans are less likely to have chronic conditions documented within the CC-MBSF as compared to those enrolled in Traditional Medicare (TM) plans.^7^ As a result, beneficiary insurance status may be associated with the under-reporting of comorbidities, a form of informative presence bias.^8^

Experts have called for improvements in the documentation of cohort selection methodologies to ensure retrospective analyses of observational data are reproducible and minimize potential bias.^9^ However, examples demonstrating the impact informative presence bias can have in studies using CMS administrative claims data remain largely absent from the literature. Using a longitudinal cohort of Medicare beneficiaries undergoing coronary artery bypass grafting (CABG), we demonstrate the impact of informative presence bias related to MA-enrollment on survival analyses.

## METHODS

### Study Design

We retrospectively reviewed CMS data from 1999 to 2019. The Baylor Scott & White Research Institute institutional review board provided approval (IRB#: 019-270, dated 8-23-2019).

A 100% sample of Medicare beneficiary hospitalizations during which CABG was performed was identified from the MedPAR file using ICD-9/10-PCS codes (**Supplemental Table 1**). Beneficiaries undergoing concomitant cardiac surgery were excluded (**Supplemental Table 2**). Demographics and social determinants of health including neighborhood disadvantage at the time of CABG were established from the MBSF. Neighborhood deprivation was estimated using the Area Deprivation Index (ADI), a validated ranking of socioeconomic contextual disadvantage derived from census data and linked to 9-digit ZIP code data available from MedPAR.^10^ Comorbidities present at the time of surgery were obtained from the CC-MBSF. Annual hospital Medicare CABG volumes were tallied and included as a variable for analysis.

The incidences of documented comorbidities among MA-enrolled and TM-enrolled beneficiaries were measured. Three survival analyses were undertaken to evaluate whether the assumption that MA-enrolled beneficiaries have adequately documented comorbidities in CC-MBSF is appropriate: (1) unweighted, (2) weighted for demographics, social determinants of health, admission urgency, and procedural characteristics but <u>not</u> comorbidities, and (3) weighted for demographics, social determinants of health, admission urgency, procedural characteristics, <u>and</u> comorbidities.

### Primary Endpoint

The primary endpoint was all-cause mortality. Vital status and date of death were determined from the MBSF.

### Statistical Analysis

Year-to-year trends in the distribution of beneficiary insurance status were evaluated using the Cochran-Armitage test. Overlap weighting (OW) adjusted for imbalances between MA-enrolled and TM-enrolled beneficiary demographics, social determinants of health, admission urgency, surgical characteristics, and, in the second weighted analysis, comorbidities (**Supplemental Methods**).^11,12^ Between-group differences were evaluated using standardized mean differences (SMD) before and after OW.^13^ Survival probabilities were estimated using the Kaplan-Meier method, before and after OW. 95% confidence intervals (CI) for weighted KM analyses and weighted median survival estimates were obtained using a non-parametric bootstrap procedure.^14^ Data management was conducted using SAS 9.4 (SAS Institute, Cary, NC). Statistical analyses were conducted with Stata software, version Stata/MP 18.0 (StataCorp, College Station, TX).

## RESULTS

3,015,066 Medicare beneficiaries underwent CABG by ICD-9/10-PCS codes from 1999-2019, of whom 2,345,476 underwent isolated-CABG and had suitable data for analysis: 354,254 (15.1%) beneficiaries were MA-enrolled and 1,991,222 (84.9%) were TM-enrolled (**Figure 1**). The annual percentage of MA-enrolled beneficiaries remained low and stable from 1999-2007 (1.1-4.4%), but after 2007 increased annually to 38.2% in 2019, (Cochran-Armitage Z-statistic 573, P<0.001, **Figure 2**).

**Figure 1.**
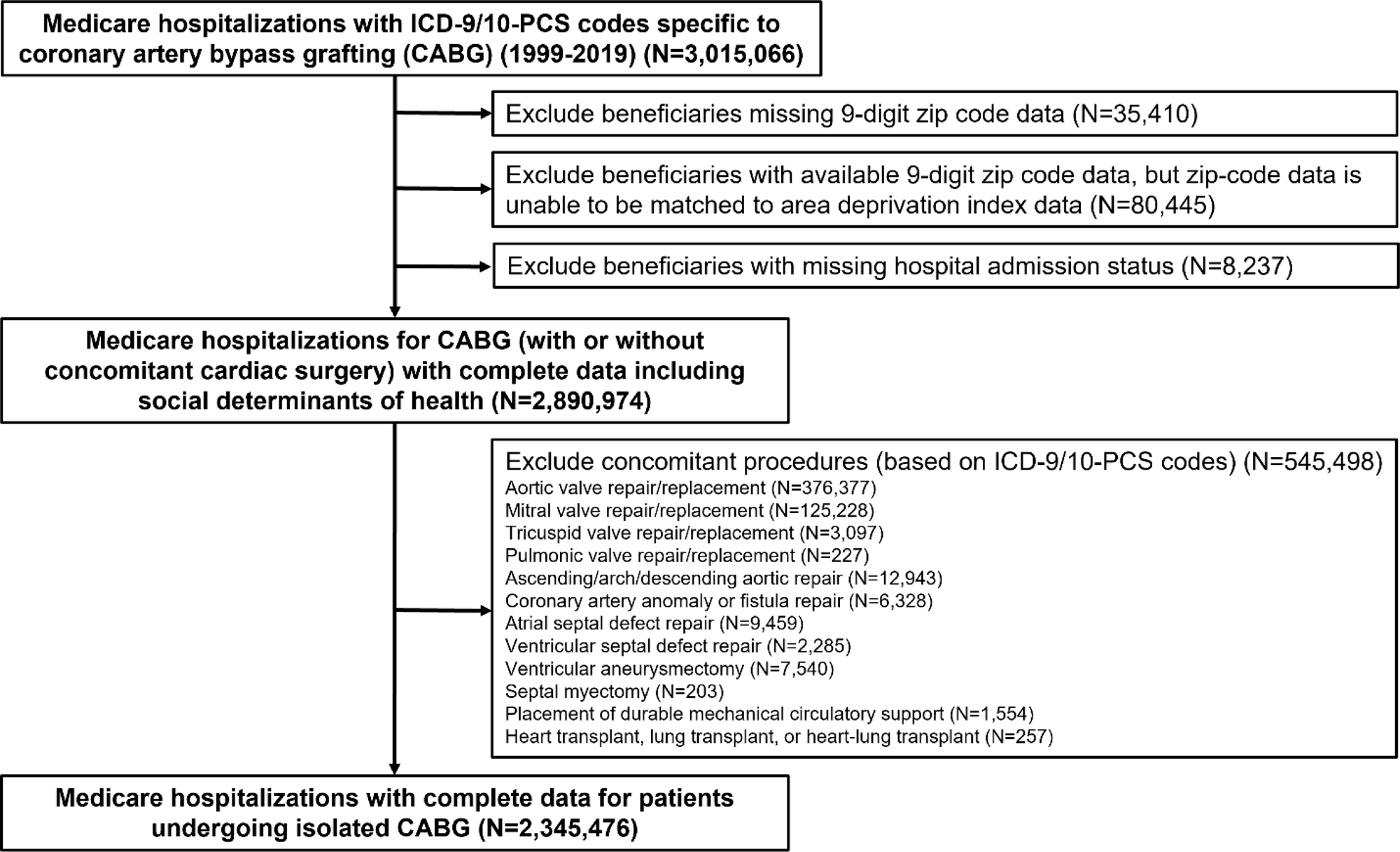
Patient selection flow diagram for the derivation of the study cohort.

**Figure 2.**
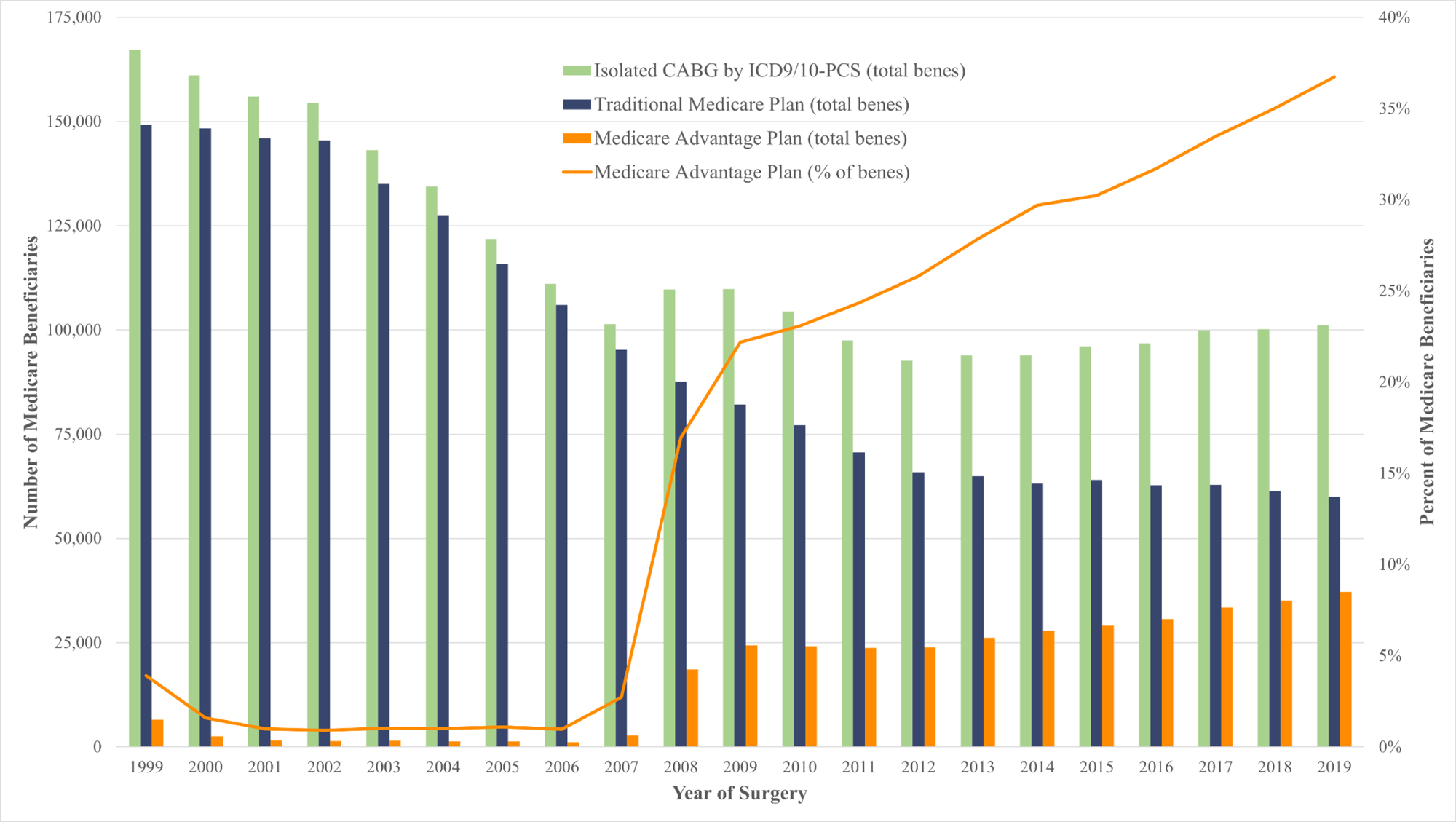
Annual Trends of Insurance Status in Medicare Beneficiaries Undergoing Isolated CABG.

Compared to TM-enrolled beneficiaries, MA-enrolled beneficiaries were older, more commonly male, less commonly white, more likely residing in the Western US, and more likely to have undergone surgery recently (**Table 1**). Neighborhood disadvantage as assessed by ADI was similar between MA-enrolled and TM-enrolled beneficiaries. The incidences of all documented preoperative comorbidities were significantly higher in TM-enrolled beneficiaries (**Table 1**). For reference, the incidence of dialysis dependence (a variable maintained separately by CMS in the MedPAR file) in MA-enrolled vs TM-enrolled beneficiaries was 2.6% vs 3.5%, SMD=0.052, yet the incidence of chronic kidney disease (as recorded in the CC-MBSF) was 10.2% vs 20.6%, SMD=0.292 in MA- vs TM-enrolled beneficiaries.

**Table 1.**
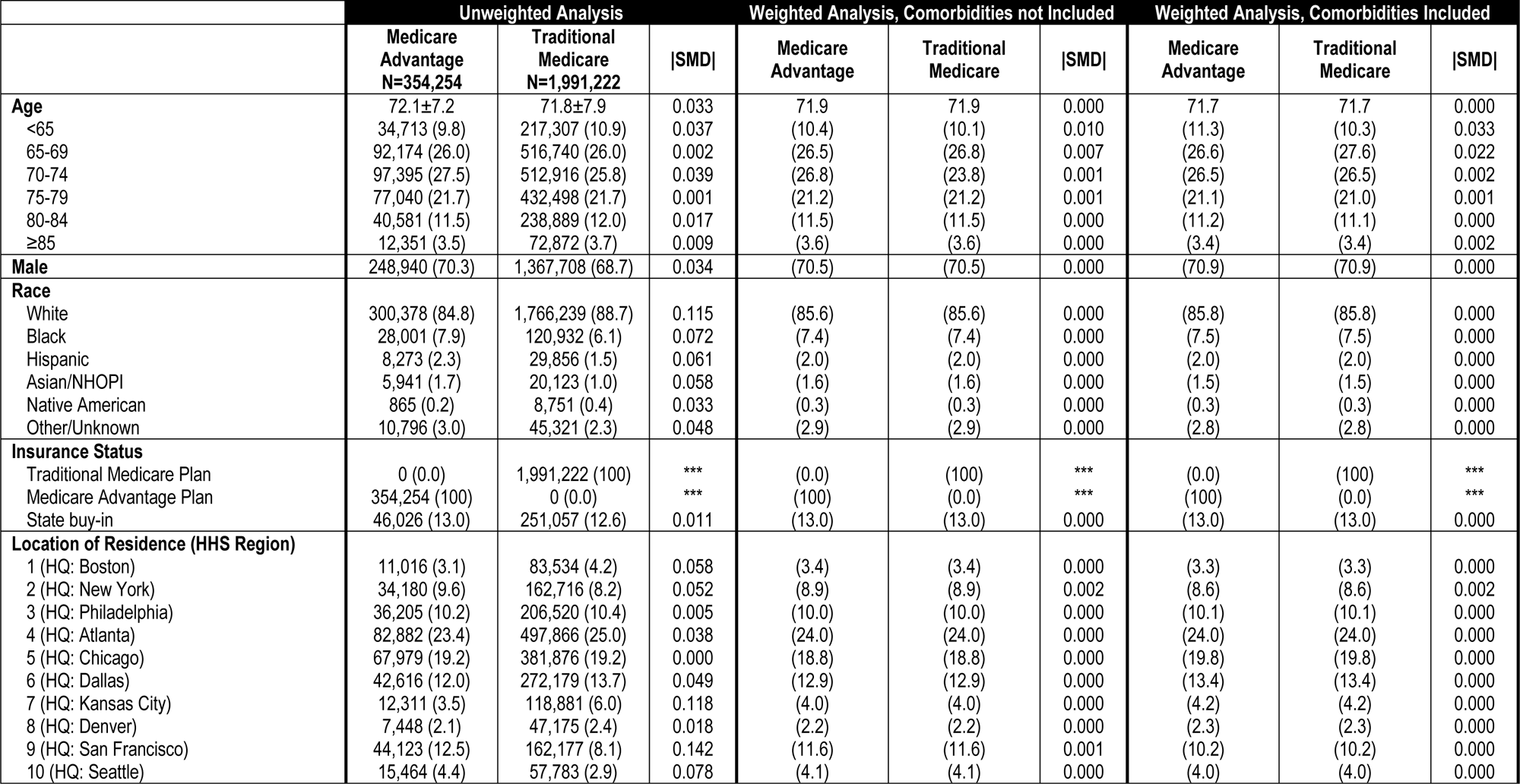
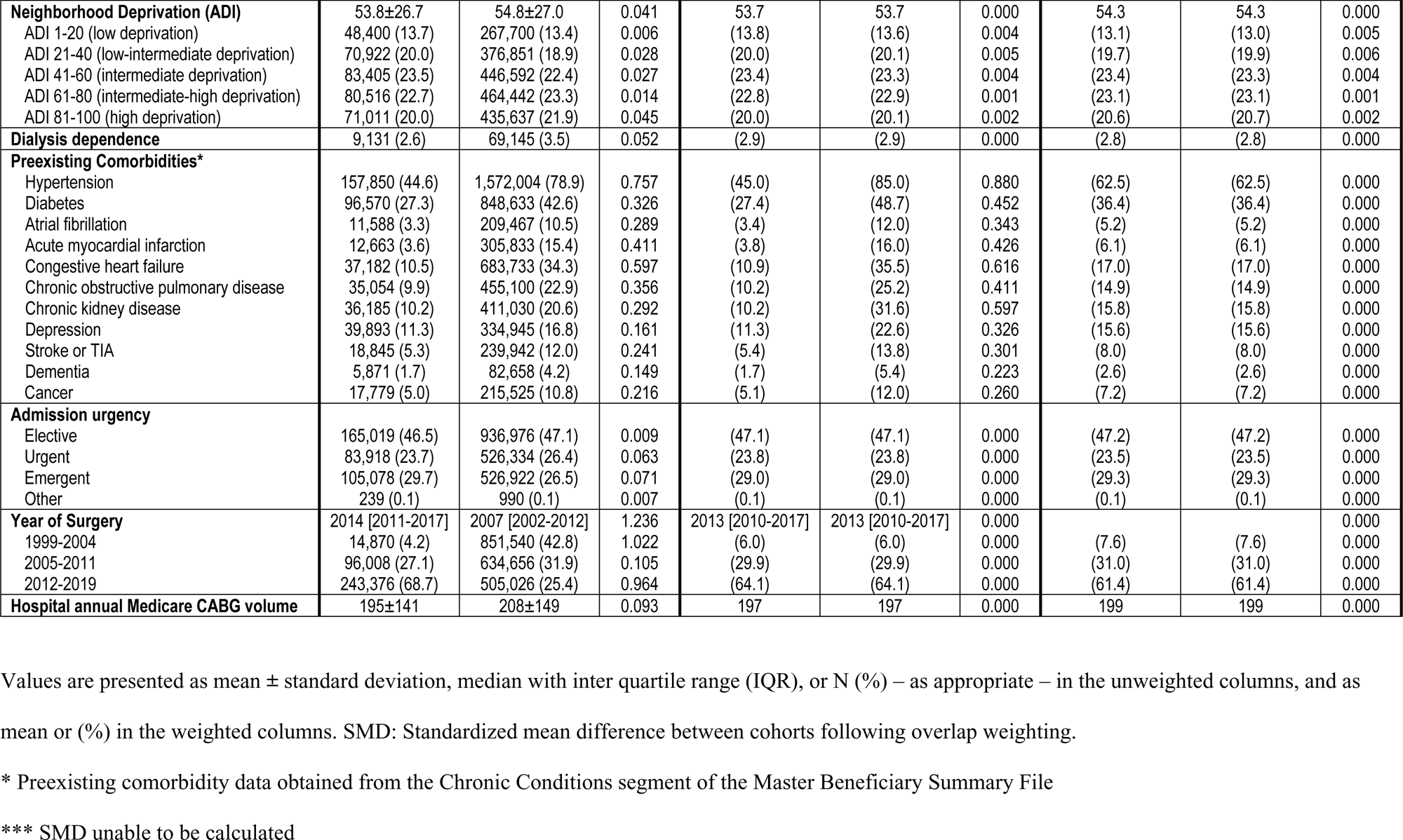
Baseline demographics, comorbidities, and procedural characteristics of all Medicare beneficiaries undergoing isolated coronary artery bypass grafting (CABG), stratified by insurance status. Distribution of these variables before and after two overlap weighting analyses are shown in three sets of columns.

|  | Unweighted Analysis |  |  | Weighted Analysis, Comorbidities not Included |  |  | Weighted Analysis, Comorbidities Included |  |  |
| --- | --- | --- | --- | --- | --- | --- | --- | --- | --- |
|  | Medicare Advantage<br>N=354,254 | Traditional Medicare<br>N=1,991,222 | \|SMD\| | Medicare Advantage | Traditional Medicare | \|SMD\| | Medicare Advantage | Traditional Medicare | \|SMD\| |
| **Age** | 72.1±7.2 | 71.8±7.9 | 0.033 | 71.9 | 71.9 | 0.000 | 71.7 | 71.7 | 0.000 |
| <65 | 34,713 (9.8) | 217,307 (10.9) | 0.037 | (10.4) | (10.1) | 0.010 | (11.3) | (10.3) | 0.033 |
| 65-69 | 92,174 (26.0) | 516,740 (26.0) | 0.002 | (26.5) | (26.8) | 0.007 | (26.6) | (27.6) | 0.022 |
| 70-74 | 97,395 (27.5) | 512,916 (25.8) | 0.039 | (26.8) | (23.8) | 0.001 | (26.5) | (26.5) | 0.002 |
| 75-79 | 77,040 (21.7) | 432,498 (21.7) | 0.001 | (21.2) | (21.2) | 0.001 | (21.1) | (21.0) | 0.001 |
| 80-84 | 40,581 (11.5) | 238,889 (12.0) | 0.017 | (11.5) | (11.5) | 0.000 | (11.2) | (11.1) | 0.000 |
| ≥85 | 12,351 (3.5) | 72,872 (3.7) | 0.009 | (3.6) | (3.6) | 0.000 | (3.4) | (3.4) | 0.002 |
| **Male** | 248,940 (70.3) | 1,367,708 (68.7) | 0.034 | (70.5) | (70.5) | 0.000 | (70.9) | (70.9) | 0.000 |
| **Race** |  |  |  |  |  |  |  |  |  |
| White | 300,378 (84.8) | 1,766,239 (88.7) | 0.115 | (85.6) | (85.6) | 0.000 | (85.8) | (85.8) | 0.000 |
| Black | 28,001 (7.9) | 120,932 (6.1) | 0.072 | (7.4) | (7.4) | 0.000 | (7.5) | (7.5) | 0.000 |
| Hispanic | 8,273 (2.3) | 29,856 (1.5) | 0.061 | (2.0) | (2.0) | 0.000 | (2.0) | (2.0) | 0.000 |
| Asian/NHOPI | 5,941 (1.7) | 20,123 (1.0) | 0.058 | (1.6) | (1.6) | 0.000 | (1.5) | (1.5) | 0.000 |
| Native American | 865 (0.2) | 8,751 (0.4) | 0.033 | (0.3) | (0.3) | 0.000 | (0.3) | (0.3) | 0.000 |
| Other/Unknown | 10,796 (3.0) | 45,321 (2.3) | 0.048 | (2.9) | (2.9) | 0.000 | (2.8) | (2.8) | 0.000 |
| **Insurance Status** |  |  |  |  |  |  |  |  |  |
| Traditional Medicare Plan | 0 (0.0) | 1,991,222 (100) | *** | (0.0) | (100) | *** | (0.0) | (100) | *** |
| Medicare Advantage Plan | 354,254 (100) | 0 (0.0) | *** | (100) | (0.0) | *** | (100) | (0.0) | *** |
| State buy-in | 46,026 (13.0) | 251,057 (12.6) | 0.011 | (13.0) | (13.0) | 0.000 | (13.0) | (13.0) | 0.000 |
| **Location of Residence (HHS Region)** |  |  |  |  |  |  |  |  |  |
| 1 (HQ: Boston) | 11,016 (3.1) | 83,534 (4.2) | 0.058 | (3.4) | (3.4) | 0.000 | (3.3) | (3.3) | 0.000 |
| 2 (HQ: New York) | 34,180 (9.6) | 162,716 (8.2) | 0.052 | (8.9) | (8.9) | 0.002 | (8.6) | (8.6) | 0.002 |
| 3 (HQ: Philadelphia) | 36,205 (10.2) | 206,520 (10.4) | 0.005 | (10.0) | (10.0) | 0.000 | (10.1) | (10.1) | 0.000 |
| 4 (HQ: Atlanta) | 82,882 (23.4) | 497,866 (25.0) | 0.038 | (24.0) | (24.0) | 0.000 | (24.0) | (24.0) | 0.000 |
| 5 (HQ: Chicago) | 67,979 (19.2) | 381,876 (19.2) | 0.000 | (18.8) | (18.8) | 0.000 | (19.8) | (19.8) | 0.000 |
| 6 (HQ: Dallas) | 42,616 (12.0) | 272,179 (13.7) | 0.049 | (12.9) | (12.9) | 0.000 | (13.4) | (13.4) | 0.000 |
| 7 (HQ: Kansas City) | 12,311 (3.5) | 118,881 (6.0) | 0.118 | (4.0) | (4.0) | 0.000 | (4.2) | (4.2) | 0.000 |
| 8 (HQ: Denver) | 7,448 (2.1) | 47,175 (2.4) | 0.018 | (2.2) | (2.2) | 0.000 | (2.3) | (2.3) | 0.000 |
| 9 (HQ: San Francisco) | 44,123 (12.5) | 162,177 (8.1) | 0.142 | (11.6) | (11.6) | 0.001 | (10.2) | (10.2) | 0.000 |
| 10 (HQ: Seattle) | 15,464 (4.4) | 57,783 (2.9) | 0.078 | (4.1) | (4.1) | 0.000 | (4.0) | (4.0) | 0.000 |
| **Neighborhood Deprivation (ADI)** | 53.8±26.7 | 54.8±27.0 | 0.041 | 53.7 | 53.7 | 0.000 | 54.3 | 54.3 | 0.000 |
| ADI 1-20 (low deprivation) | 48,400 (13.7) | 267,700 (13.4) | 0.006 | (13.8) | (13.6) | 0.004 | (13.1) | (13.0) | 0.005 |
| ADI 21-40 (low-intermediate deprivation) | 70,922 (20.0) | 376,851 (18.9) | 0.028 | (20.0) | (20.1) | 0.005 | (19.7) | (19.9) | 0.006 |
| ADI 41-60 (intermediate deprivation) | 83,405 (23.5) | 446,592 (22.4) | 0.027 | (23.4) | (23.3) | 0.004 | (23.4) | (23.3) | 0.004 |
| ADI 61-80 (intermediate-high deprivation) | 80,516 (22.7) | 464,442 (23.3) | 0.014 | (22.8) | (22.9) | 0.001 | (23.1) | (23.1) | 0.001 |
| ADI 81-100 (high deprivation) | 71,011 (20.0) | 435,637 (21.9) | 0.045 | (20.0) | (20.1) | 0.002 | (20.6) | (20.7) | 0.002 |
| **Dialysis dependence** | 9,131 (2.6) | 69,145 (3.5) | 0.052 | (2.9) | (2.9) | 0.000 | (2.8) | (2.8) | 0.000 |
| **Preexisting Comorbidities\*** |  |  |  |  |  |  |  |  |  |
| Hypertension | 157,850 (44.6) | 1,572,004 (78.9) | 0.757 | (45.0) | (85.0) | 0.880 | (62.5) | (62.5) | 0.000 |
| Diabetes | 96,570 (27.3) | 848,633 (42.6) | 0.326 | (27.4) | (48.7) | 0.452 | (36.4) | (36.4) | 0.000 |
| Atrial fibrillation | 11,588 (3.3) | 209,467 (10.5) | 0.289 | (3.4) | (12.0) | 0.343 | (5.2) | (5.2) | 0.000 |
| Acute myocardial infarction | 12,663 (3.6) | 305,833 (15.4) | 0.411 | (3.8) | (16.0) | 0.426 | (6.1) | (6.1) | 0.000 |
| Congestive heart failure | 37,182 (10.5) | 683,733 (34.3) | 0.597 | (10.9) | (35.5) | 0.616 | (17.0) | (17.0) | 0.000 |
| Chronic obstructive pulmonary disease | 35,054 (9.9) | 455,100 (22.9) | 0.356 | (10.2) | (25.2) | 0.411 | (14.9) | (14.9) | 0.000 |
| Chronic kidney disease | 36,185 (10.2) | 411,030 (20.6) | 0.292 | (10.2) | (31.6) | 0.597 | (15.8) | (15.8) | 0.000 |
| Depression | 39,893 (11.3) | 334,945 (16.8) | 0.161 | (11.3) | (22.6) | 0.326 | (15.6) | (15.6) | 0.000 |
| Stroke or TIA | 18,845 (5.3) | 239,942 (12.0) | 0.241 | (5.4) | (13.8) | 0.301 | (8.0) | (8.0) | 0.000 |
| Dementia | 5,871 (1.7) | 82,658 (4.2) | 0.149 | (1.7) | (5.4) | 0.223 | (2.6) | (2.6) | 0.000 |
| Cancer | 17,779 (5.0) | 215,525 (10.8) | 0.216 | (5.1) | (12.0) | 0.260 | (7.2) | (7.2) | 0.000 |
| **Admission urgency** |  |  |  |  |  |  |  |  |  |
| Elective | 165,019 (46.5) | 936,976 (47.1) | 0.009 | (47.1) | (47.1) | 0.000 | (47.2) | (47.2) | 0.000 |
| Urgent | 83,918 (23.7) | 526,334 (26.4) | 0.063 | (23.8) | (23.8) | 0.000 | (23.5) | (23.5) | 0.000 |
| Emergent | 105,078 (29.7) | 526,922 (26.5) | 0.071 | (29.0) | (29.0) | 0.000 | (29.3) | (29.3) | 0.000 |
| Other | 239 (0.1) | 990 (0.1) | 0.007 | (0.1) | (0.1) | 0.000 | (0.1) | (0.1) | 0.000 |
| **Year of Surgery** | 2014 [2011-2017] | 2007 [2002-2012] | 1.236 | 2013 [2010-2017] | 2013 [2010-2017] | 0.000 |  |  | 0.000 |
| 1999-2004 | 14,870 (4.2) | 851,540 (42.8) | 1.022 | (6.0) | (6.0) | 0.000 | (7.6) | (7.6) | 0.000 |
| 2005-2011 | 96,008 (27.1) | 634,656 (31.9) | 0.105 | (29.9) | (29.9) | 0.000 | (31.0) | (31.0) | 0.000 |
| 2012-2019 | 243,376 (68.7) | 505,026 (25.4) | 0.964 | (64.1) | (64.1) | 0.000 | (61.4) | (61.4) | 0.000 |
| **Hospital annual Medicare CABG volume** | 195±141 | 208±149 | 0.093 | 197 | 197 | 0.000 | 199 | 199 | 0.000 |

Values are presented as mean ± standard deviation, median with inter quartile range (IQR), or N (%) – as appropriate – in the unweighted columns, and as mean or (%) in the weighted columns. SMD: Standardized mean difference between cohorts following overlap weighting.
\* Preexisting comorbidity data obtained from the Chronic Conditions segment of the Master Beneficiary Summary File
\*\*\* SMD unable to be calculated

Median follow-up was 12.15 [95% CI: 6.17-17.08] years. Unweighted survival analysis suggested an early survival advantage for MA-enrolled beneficiaries but a late survival advantage for TM-enrolled beneficiaries, with similar median survival: median survival difference 8 [-12,28] days (10.02 [9.96,10.07] vs 10.00 [9.98,10.01] years), respectively **(Figure 3a)**. After applying OW to adjust for potential confounding variables aside from comorbidities, we no longer noted the early survival advantage for MA-enrolled beneficiaries, although a slight late survival advantage in TM-enrolled beneficiaries did persist (**Figure 3b**). Weighted median survival was similar between MA-enrolled and TM-enrolled beneficiaries: median survival difference -2 [-28,24] days (10.00 [9.95,10.06] vs 10.01 [9.98,10.04] years). To demonstrate the effect of informative presence bias, OW was repeated including comorbidities. This repeated weighted analysis exhibited dramatically reduced survival in MA-enrolled vs TM-enrolled beneficiaries, median survival difference of -604 [-626,-575] days (9.78 [9.73,9.83] vs 11.44 [11.41,11.47] years) **(Figure 3c)**.

**Figure 3.**
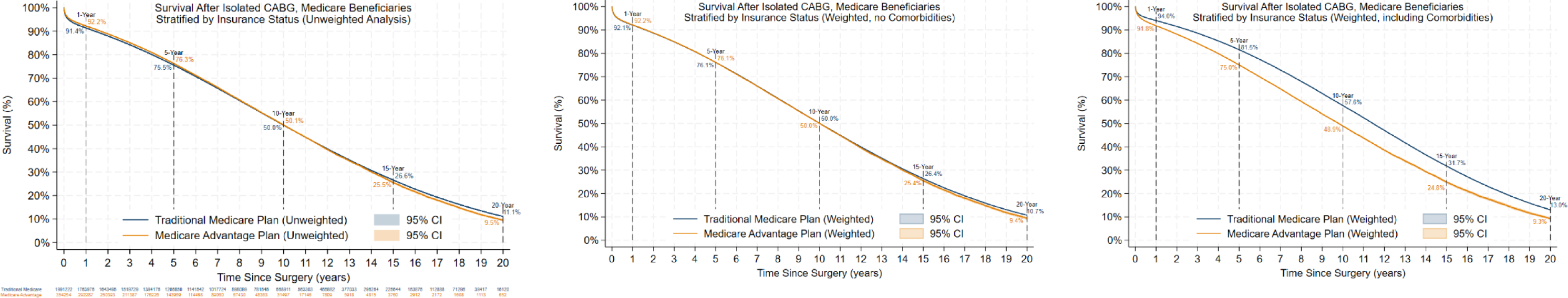
Post-CABG survival estimates for Medicare beneficiaries, stratified by insurance status (Traditional Medicare Plan vs Medicare Advantage Plan). Unweighted (Figure 3a), weighted by non-comorbidity covariates (Figure 3b), and weighted including comorbidity covariates (Figure 3c) survival analyses shown.

## DISCUSSION

This analysis evaluated the incidence of MA and TM enrollment, and long-term survival, among Medicare beneficiaries undergoing isolated-CABG from 1999-2019. We highlight the increasing number of beneficiaries enrolled in MA over time, findings concordant with prior studies.^15^ We demonstrate that the incidence of comorbidities documented in the CC-MBSF are dramatically lower for MA-enrolled beneficiaries as compared to TM-enrolled beneficiaries, likely related to the under-reporting of comorbidity claims data in MA-enrolled beneficiaries that has been previously described.^7^ Despite differences in documented comorbidities, both unweighted and weighted (without comorbidities) survival were similar among MA-enrolled and TM-enrolled beneficiaries. This finding of similar post-CABG survival between MA-enrolled and TM-enrolled beneficiaries is concordant with the hypothesis that MA-enrolled and TM-enrolled beneficiaries share a similar incidence of comorbidities, despite the CC-MBSF data suggesting otherwise.

We suggest that the median survival advantage of 604 days for TM-enrolled over MA-enrolled beneficiaries noted in our weighted analysis with comorbidities helps to quantify the amount of bias to which comparative analyses are susceptible if they include both MA-enrolled and TM-enrolled beneficiaries and their documented comorbidities. We recommend that comparative analyses using CMS comorbidity data should be limited to TM-enrolled beneficiaries to avoid introducing informative presence bias. We echo the sentiments of other research groups that releasing claims data for MA-enrolled beneficiaries will mitigate the dwindling cohort of TM-enrolled Medicare beneficiaries with adequate CC-MBSF data for robust comparative analyses.^16^

## Data Availability

Due to a data use agreement with CMS, we are unable to provide the data referenced in our study.

## ACKNOWLEDGEMENTS

We are especially grateful to Alessandro Gasparini (Red Door Analytics), for his helpful feedback regarding our statistical methodologies.

## ABBREVIATION LIST

ADI: area deprivation index
CABG: coronary artery bypass grafting
CC-MBSF: chronic conditions segment of the master beneficiary summary file
CM: clinical modification
CMS: centers for Medicare and Medicaid services
ICD-9/10: international classification of diseases, ninth and tenth revision
MA: Medicare advantage insurance plan
MBSF: Master Beneficiary Summary File
MedPAR: Medicare provider analysis and review file
OW: overlap weighting
PCS: procedural coding system
TM: traditional Medicare insurance plan

## REFERENCES

1 Mues KE, Liede A, Liu J, et al. Use of the Medicare database in epidemiologic and health services research: A valuable source of real-world evidence on the older and disabled populations in the US. Clin Epidemiol 2017;9(9):276–177.

2 Cartwright, DJ. ICD-9-CM to ICD-10-CM codes: What? Why? How?. Adv Wound Care (New Rochelle) 2013;2(10):588–592.

3 Goodman AR, Posner SF, Huang ES, Parekh AK, Koh HK. Defining and measuring chronic conditions: Imperatives for research, policy, program, and practice. Prev Chronic Dis 2013;10:E66.

4 Hall M, Attard TM, Berry JG. Improving cohort definitions in research using hospital administrative databases—Do we need guidelines?. JAMA Pediatr 2022;176(6):539–540.

5 Khera R, Dorsey KB, Krumholz HM. Transition to the ICD-10 in the United States: an emerging data chasm. JAMA 2018;320(2):133–134.

6 Weeks WB, Huynh G, Cao ST, Smith J, Weinstein JN. Assessment of year-to-year patient-specific comorbid conditions reported in the Medicare Chronic Conditions Data Warehouse. JAMA Netw Open 2020;3(10):e2018176.

7 Meyers DJ, Johnston KJ. The growing importance of Medicare Advantage in health policy and health. JAMA Health Forum 2021;2(3):e210235.

8 Sisk R, Lin L, Sperrin M, et al. Informative presence and observation in routine health data: A review of methodology for clinical risk prediction. J Am Med Inform Assoc 2021;28(1):155–166.

9 Benchimol EI, Smeeth L, Guttmann A, et al, The REporting of studies Conducted using Observational Routinely-collected health Data (RECORD) Statement. PLoS Med 2015;12(10):e1001885.

10 Kind AJH, Buckingham WR. Making neighborhood-disadvantage metrics accessible the Neighborhood Atlas. N Engl J Med. 2018;378:2456–8

11 Cheng C, Li F, Thomas LE, Li FF. Addressing extreme propensity scores in estimating counterfactual survival functions via the overlap weights. Am J Epidemiol 2022;191(6):1140–1151

12 Thomas LE, Li F, Penicina MJ. Overlap weighting: A propensity score method that mimics attributes of a randomized clinical trial. JAMA 2020;323(23):2417–2418.

13 Austin PC. Balance diagnostics for comparing the distribution of baseline covariates between treatment groups in propensity-score matched samples. Stat Med 2009;28(25):3083–3107.

14 Rogawski ET, Westreich DJ, Kang G, Ward HD, Cole SR. Brief report: Estimating differences and ratios in median times to event. Epidemiology 2016;27(6):848–51.

15 Neuman P, Jacobson GA. Medicare Advantage checkup. N Eng J Med 2018;379(22):2163–2172.

16 Brennan N, Ornstein C, Frakt AB. Time to release Medicare Advantage claims data. JAMA 2018;319(10):975–976.

